# Kangaroo Mother Care coverage and quality in 66 neonatal units implementing with NEST360 across Kenya, Malawi, Nigeria, and Tanzania

**DOI:** 10.64898/2026.11.05.26367332

**Authors:** Eleanor Reynolds, Julius Thomas, Edith Wathira Gicheha, Kristina Shemwell, Kylie Dougherty, Cathy Chambani Magombo, Dolphine Mochache, Josephat Cyprian Mutakyamilwa, Grace Tahuna Soko, Afeez Adedoyin Idowu, Domitilla Muinde, Mary Waiyego, Olukemi Tongo, Nahya Salim, Josephine Shabani, Veronica Chinyere Ezeaka, William Macharia, Msandeni Chiume, Kondwani Kawaza, Samuel Kalembera Ngwala, George Okello, Opeyemi Odedere, Catherine Paul, Lucas Malla, John Wainaina, Zillah Maria Oden, Rebecca Richards-Kortum, Lisa R Hirschhorn, Joy E Lawn, Natasha R Rhoda, Sarah Murless-Collins, Christine A Bohne

**Affiliations:** London School of Hygiene and Tropical Medicine; Rice University; 1. Nationwide Children’s Hospital, Center for Nursing Excellence, Columbus, USA 2. Abigail Wexner Research Institute, Center for Child Health Outcomes Research, Columbus, USA; Aga Khan University; Ifakara Health Institute; Kamuzu University of Health Sciences; APIN Public Health Initiatives, Abuja, Nigeria; Aga Khan University, Nairobi, Kenya; Kenyatta National Hospital; University of Ibadan; 1. Muhimbili University of Health and Allied Sciences (MUHAS), Department of Pediatrics and Child Health 2. Ifakara Health Institute (IHI); 1. Ifakara Health Institute 2. Faculty of Epidemiology and Population Health, London School of Hygiene & Tropical Medicine, London, UK; University of Lagos; Aga Khan University of Nairobi; 1. Ministry of Health, Reproductive Health Department 2. Kamuzu University of Health Sciences; Department of Paediatrics and Child Health, School of Medicine and Oral Health, Kamuzu University of Health Sciences, Blantyre, Malawi; Rice360, Rice University; Rice University, NEST360; Department of Medical Social Sciences and Havey Institute for Global Health, Feinberg School of Medicine, Northwestern University

**Keywords:** Newborn, Low- and Middle-Income Countries, Inpatient Care, Kangaroo Mother Care, Small and Sick Newborn Care

## Abstract

**Background:** Kangaroo Mother Care (KMC) reduces neonatal mortality and morbidity among preterm and low birthweight (LBW) infants, but implementation and quality vary across sub-Saharan African (SSA) hospitals. This study reports KMC coverage and quality of care for preterm and LBW neonates admitted to hospitals in the NEST360 Alliance (Kenya, Malawi, Tanzania, Nigeria) and examines the association between KMC provision and survival to discharge.

**Methods:** We conducted a cross-sectional study using routinely collected neonatal inpatient data from hospitals participating in the NEST360 Alliance. Newborns weighing under 2500g were eligible for KMC and included in coverage and quality analyses (January 2021-December 2024). Quality-of-care cascades quantified attrition along the KMC care pathway from eligibility to successful delivery, defining quality as the proportion of eligible neonates reaching each successive step. Mixed-effects logistic regression assessed the association between KMC and survival at discharge, including variation by illness severity and, among KMC recipients, by timing of initiation across severity and birthweight strata.

**Results:** Of 375,238 neonatal admissions across 66 hospitals, 125,844 neonates were eligible for KMC and had KMC documentation available. Among these, 64,619 (51.3%) received KMC. All four countries showed improved coverage over the study period, with the largest absolute increases in Nigeria (27.6%) and Tanzania (27.4%). Excluding Kenya, where KMC timing data were unavailable, 92.1% of neonates started on KMC had a documented start date; of these 44,264 neonates, 12,934 (29.2%) were initiated on KMC on the first day of life. The quality cascade showed eligible neonates were lost at each step, from eligibility through to documented, timely, and sustained KMC. Among neonates eligible for both KMC and CPAP, 39.4% of those started on KMC also received documented CPAP at some point during admission. After adjustment for patient-level factors, receiving KMC was associated with improved survival across all severity groups, with the greatest absolute benefit among the most critically ill neonates (adjusted predicted survival 7% vs 80% without and with KMC, respectively).

**Conclusion:** KMC coverage improved substantially across NEST360-implementing hospitals between 2021 and 2024, demonstrating progress in implementation. However, important gaps remain in the quality and timeliness of KMC delivery. More investment in high-quality KMC implementation, supported by robust monitoring systems linked with quality improvement, are essential to improve outcomes for small and sick newborns across sub-Saharan Africa.

**KEY FINDINGS:** 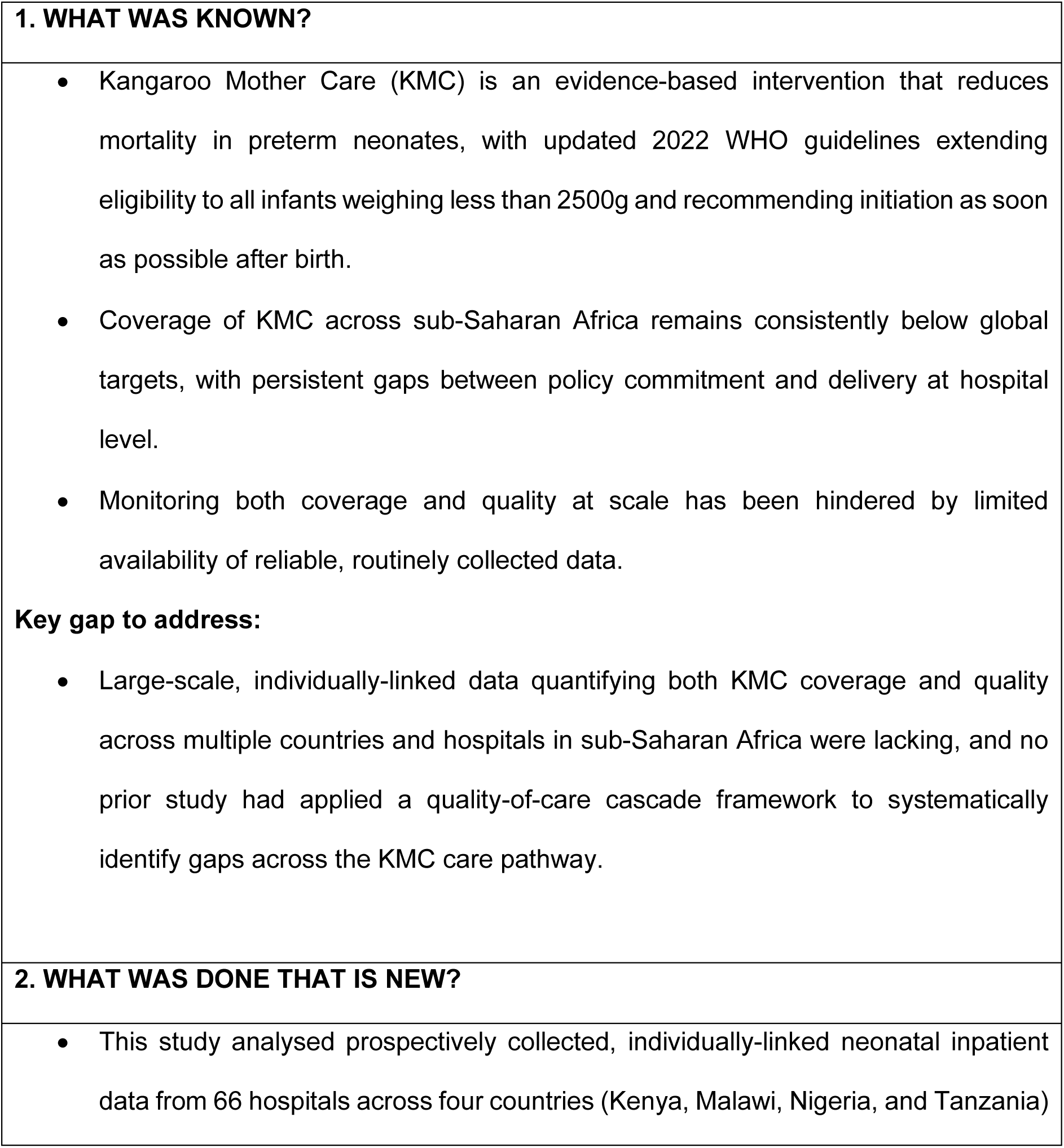

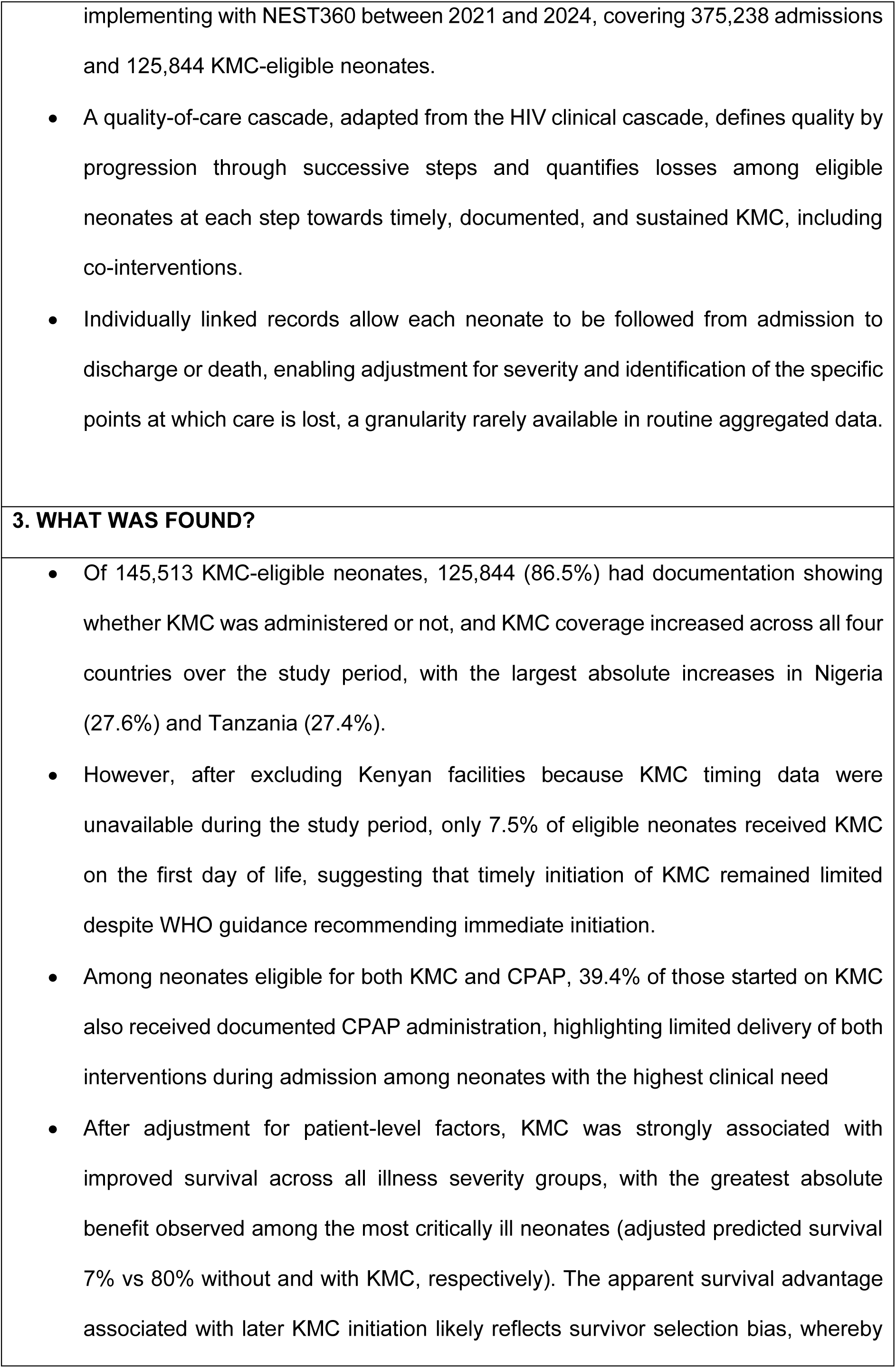

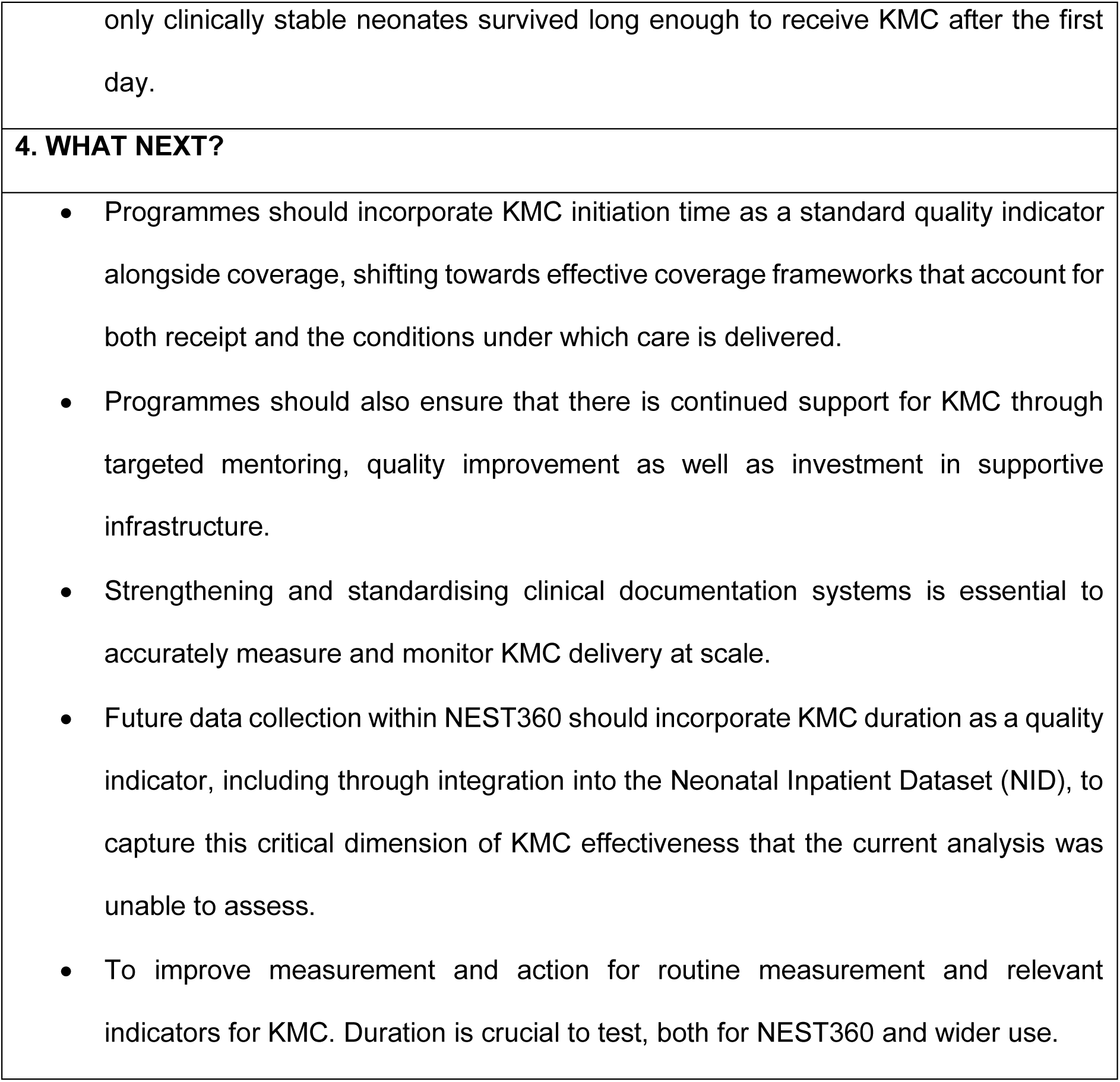

## BACKGROUND

Preterm birth remains the leading cause of death among children under five years of age worldwide [1]. In 2024, an estimated 4.9 million children died before the age of five, including 2.3 million newborns - nearly half of all under-five deaths [1]. SDG target 3.2 aims to reduce neonatal mortality to ≤12 deaths per 1,000 live births by 2030; however, current trajectories indicate that 65 countries are at risk of missing this benchmark [1] indicating that substantial improvements are required in the care of preterm neonates, where low birthweight (LBW) acts as a common marker. KMC is a proven, high-impact, intervention that significantly reduces mortality among infants with most evidence focused on <2000g [2], and randomised controlled trials demonstrating substantial reductions in mortality compared with conventional care [3,4]. Short-term benefits include reduced risk of hypothermia, severe infection, and improved exclusive breastfeeding rates [5]. Longer-term benefits extend to improved neurodevelopmental and behavioural outcomes, but have more limited data [6,7]. Beyond clinical gains, one studies has shown KMC is cost-effective even when initiated prior to clinical stabilisation [8], and empowers mothers as central providers of newborn care, with evidence of improved maternal wellbeing and mother-infant attachment [9]. Modelling studies further estimate that universal adoption of KMC within hospitals could prevent a large number of newborn deaths annually [10]. Reflecting this evidence, WHO guidance, which previously limited KMC to clinically stable newborns weighing ≤2000g [2], was updated in 2022 to all infants weighing <2500 g, with KMC advised to commence as soon as possible after birth [11].

Despite this strong evidence base and guidance, implementation is behind global ambition. The Every Newborn Action Plan (ENAP) set a target of KMC coverage of 75% of eligible newborns weighing less than 2000g by 2025 [12], and its successor, the Every Woman Every Newborn Everywhere (EWENE) framework [13], identifies KMC as one of four priority life-saving interventions for small and sick newborns. EWENE’s target is for functional newborn care units with the space, staffing, equipment and commodities to deliver care continuously, to be operational in in at least 80% of districts in every country by 2025. As of 2023, only 32% of reporting countries had met this threshold.

Within facilities, many lack designated KMC units or have insufficient capacity to meet demand, with shortages of infrastructure, equipment, and trained staff among the most consistently reported barriers [14,15]. A large proportion of newborns eligible for KMC will also require respiratory support [16]. Because the updated guidance recommends immediate KMC, these interventions will often need to be delivered concurrently, placing significant demands on hospital capacity, equipment, and workforce that many settings across sub-Saharan Africa are currently unable to meet [17]. Facility readiness assessments across sub-Saharan Africa have consistently identified family-centred care, including KMC, among the most poorly resourced domains of neonatal care [18,19].

Coverage is also an incomplete measure of programme success; quality of care must also be considered [3]. Binary coverage indicators capture neither timing of initiation nor daily duration, both of which affect outcomes: initiation within 24 hours of birth, has been shown to reduce neonatal mortality relative to later initiation, and at least 8 hours of daily skin to skin contact is recommended [5]. Monitoring both coverage and quality at scale has been further hindered by limited availability of reliable, routinely collected national data [20]. Consequently, despite increasing implementation of KMC programmes across sub-Saharan Africa, few multi-country studies have simultaneously evaluated coverage, quality of implementation, and survival outcomes using routine inpatient datasets at scale.

Newborn Essential Solutions and Technologies (NEST360) is an international alliance of clinical, biomedical, implementation science, and public health experts working with five African governments (Kenya, Malawi, Nigeria, Tanzania, and more recently, Ethiopia) to strengthen level-2 newborn care. NEST360 supports hospitals through an integrated package of appropriate medical technologies, biomedical engineering, leadership engagement, and strengthened routine health information systems, as well as clinical mentorship, quality improvement coaching, infrastructure support and facilitating exchange visits to high performing sites. These complementary interventions aim to improve the quality and delivery of evidence-based newborn care, including KMC [21].

### Aim and objectives

This paper is part of a supplement reporting findings and learnings from the NEST360 Alliance. We conducted a descriptive analysis of the KMC coverage, quality, and outcomes for LBW neonates across 66 neonatal units in four African countries implementing with NEST360: Kenya, Malawi, Nigeria, and Tanzania. Data on KMC coverage and quality in routine care in sub-Saharan Africa are limited. This study addresses this gap using individually linked records and a quality-of-care cascade to identify where neonates are lost along the KMC pathway.

Our objectives were:

1. **KMC Coverage:** Describe KMC coverage over time at country population-level for admitted neonates eligible to receive KMC using aggregate data.
2. **KMC quality:** Describe quality-of-care cascades including key quality indicators for eligible neonates receiving KMC using individual-level data.
3. **Outcomes for KMC:** To evaluate the association between KMC and neonatal survival at discharge, and to further assess whether this association varied by illness severity, birthweight, and timing of KMC initiation after accounting for individual- and clustering within hospitals.

## METHODS

### Overview

This study employed a multi-site, cross-sectional observational design using secondary data analysis. As part of the NEST360 program, routine data on all neonatal admissions were collected across participating hospitals for the analysis period spanning January 2021 to December 2024. The study is reported in accordance with the Strengthening the Reporting of Observational Studies in Epidemiology (STROBE) guidelines [22] (see Appendix 1).

### Study Setting

NEST360, established in 2019, has progressively expanded its implementation across 66 newborn units in level 2 hospitals. The analysis is restricted to hospitals collecting data starting in 2021. Following implementation, prospective data collection for all newborn admissions has been maintained at participating sites.

### Data sources

The Neonatal Inpatient Dataset (NID) [23] was co-designed with African governments to be a parsimonious individual dataset routinely collected for every newborn admitted to a neonatal ward and used for quality of care improvement on that ward. The NID comprises a set of ∼55 variables capturing neonatal outcomes, admission details and variables to describe high-impact care. Trained clerks’ abstract information from medical records and input data into REDCap, a secure, web-based platform.

The NID functions as the principal data source in implementing hospitals in Malawi (nationally), Tanzania, and Nigeria. In Kenya, however, the NID variables are pulled from a research-focused larger dataset, the national Clinical Information Network (CIN) [24] resulting in alternative coding structures for certain variables.

### Methods by objectives Objective 1: KMC Coverage

In alignment with national standards and current evidence, NEST 360 implementing hospitals aim to initiate KMC for infants born with a birthweight of <2500g. Coverage was stratified by weight categories (<1000g, 1000-1999g, 2000-2499g) and were reported as yearly proportions on a country population-level.

### Objective 2: KMC Quality

Individual-level data were aggregated by country to create quality-of-care cascades for neonates who received KMC by adapting the HIV clinical cascade framework established for HIV treatment by the World Health Organisation [25]. Reproductive, Maternal, Newborn, Child and Adolescent Health (RMNCAH) coverage cascade literature around service contact and coverage also informed cascade development [26,27].

Clinical quality indicators were selected based on key domains of care for small and sick newborns eligible for KMC with established relevance to neonatal survival, including the timing and continuity of KMC, breastmilk feeding practices, and administration of continuous positive airway pressure (CPAP) [11]. Timeliness of KMC was how soon in days after birth KMC was commenced, consistent with current WHO recommendations for immediate KMC for preterm and low-birthweight infants, irrespective of clinical stability [11,28].

To evaluate quality and continuity of care across these domains, three related care cascades were developed. Kenyan facilities were excluded from all cascade analyses because KMC timing data were unavailable for the study period; breastmilk timing data required for the KMC and nutrition cascade were also unavailable. For the KMC care cascade, neonates eligible for KMC were defined as all newborns admitted during the study period with birthweight <2500g. The cascade assessed, in sequence, documentation of KMC at any point during admission, initiation of KMC, initiation on the day of birth, and continuation of KMC from that first day through to discharge. Building on this cascade, the KMC and nutrition care cascade evaluated whether neonates initiated on KMC additionally received exclusive breastmilk feeding on the same day through to discharge. The KMC and CPAP care cascade further assessed integration of respiratory support among neonates eligible for both KMC and CPAP who received KMC and CPAP at any time during admission. We did not assess whether KMC and CPAP were administered concurrently. CPAP eligibility was defined as birthweight 1000– 1499g, or birthweight 1500–1999g with respiratory distress and hypoxia, in line with clinical criteria applied with NEST360-supported hospitals that is consistent with current evidence [29].

### Objective 3: Association between KMC and Survival at Discharge

To evaluate the association between KMC and survival at discharge from the neonatal unit, we conducted both primary and secondary multilevel analyses. The primary analysis assessed KMC receipt, while the secondary analysis examined the timing of KMC initiation among neonates who received KMC. Both analyses accounted for baseline illness severity and clustering of neonates within hospitals.

### Neonatal Severity Score Development

The neonatal severity score was constructed using admission characteristics known to predict neonatal mortality, including gestational age, oxygen saturation at admission, and admission temperature, drawing on established neonatal mortality risk scores developed for low-resource settings [30]. Each variable was categorized into clinically meaningful severity strata and assigned ordinal scores reflecting increasing physiological instability. Individual component scores were summed to generate a composite severity score, which was subsequently categorized into mild (3-4), moderate (5-6), and severe (7-9) illness groups (Appendix 7).

The scoring framework was developed and piloted using neonatal inpatient data to assess its ability to predict survival at discharge. In the previously published CPAP analysis [40], the mixed-effects survival model incorporating clinical severity demonstrated good discrimination (AUC = 0.74). The severity score was subsequently applied in the present study to account for baseline illness severity when examining the association between KMC receipt and survival at discharge.

#### a) Primary Survival Analysis

The association between KMC receipt and survival at discharge was assessed using mixed-effects logistic regression, with survival modeled as a binary outcome. The model included KMC coverage (receiving any KMC), baseline illness severity, sex, and birth location as fixed effects, with facility included as a random intercept to account for clustering. An interaction between KMC receipt and illness severity was included to assess whether the association with survival varied by baseline severity. The contribution of the interaction was evaluated using a likelihood ratio test comparing models with and without the interaction term. Adjusted predicted probabilities of survival and corresponding 95% confidence intervals were estimated across severity categories using marginal effects.

#### b) Secondary Analysis: Timing of KMC Initiation

Among neonates who received KMC, a secondary mixed-effects logistic regression analysis assessed the association between timing of KMC initiation and survival at discharge. Kenya was excluded from this analysis because KMC initiation timing was not routinely collected. Neonates from other countries with missing or undocumented KMC initiation timing were also excluded. Among eligible KMC recipients with documented initiation timing, timing of KMC initiation was categorized as same-day initiation (≤24 hours), early initiation (1–3 days), delayed initiation (3–7 days), and late initiation (>7 days). The model additionally adjusted for illness severity category, birthweight category, sex, and birth location, with hospital included as a random intercept.

### Statistical Analysis

All analyses were conducted in R version 4.2.2. Mixed-effects logistic regression models were fitted using the lme4 package, with hospital included as a random intercept to account for clustering of neonates within hospitals. Adjusted predicted probabilities and marginal effects were estimated using the emmeans package in R. Model discrimination was assessed using ROC curves and AUC statistics. Statistical significance was evaluated using two-sided tests with an alpha level of 0.05.

## RESULTS

### Overview

Between 1 January 2021 and 31 December 2024, 375,238 newborns were admitted across 66 neonatal units in the four countries (Malawi = 37, Kenya = 13, Nigeria = 9, Tanzania = 7). Malawi is the only country-wide scale which accounts for the disproportionate number of sites in this country. Of these newborns, 145,513 met criteria for KMC eligibility, of whom 125,844 (86.5%) had KMC documentation (administered or not administered), as shown in Figure 1 (Malawi: n = 68,274; Kenya: n = 37,282; Tanzania: n = 17,613; Nigeria: n = 2,675). Newborn characteristics across each country are described in Appendix 2.

**Figure 1:**
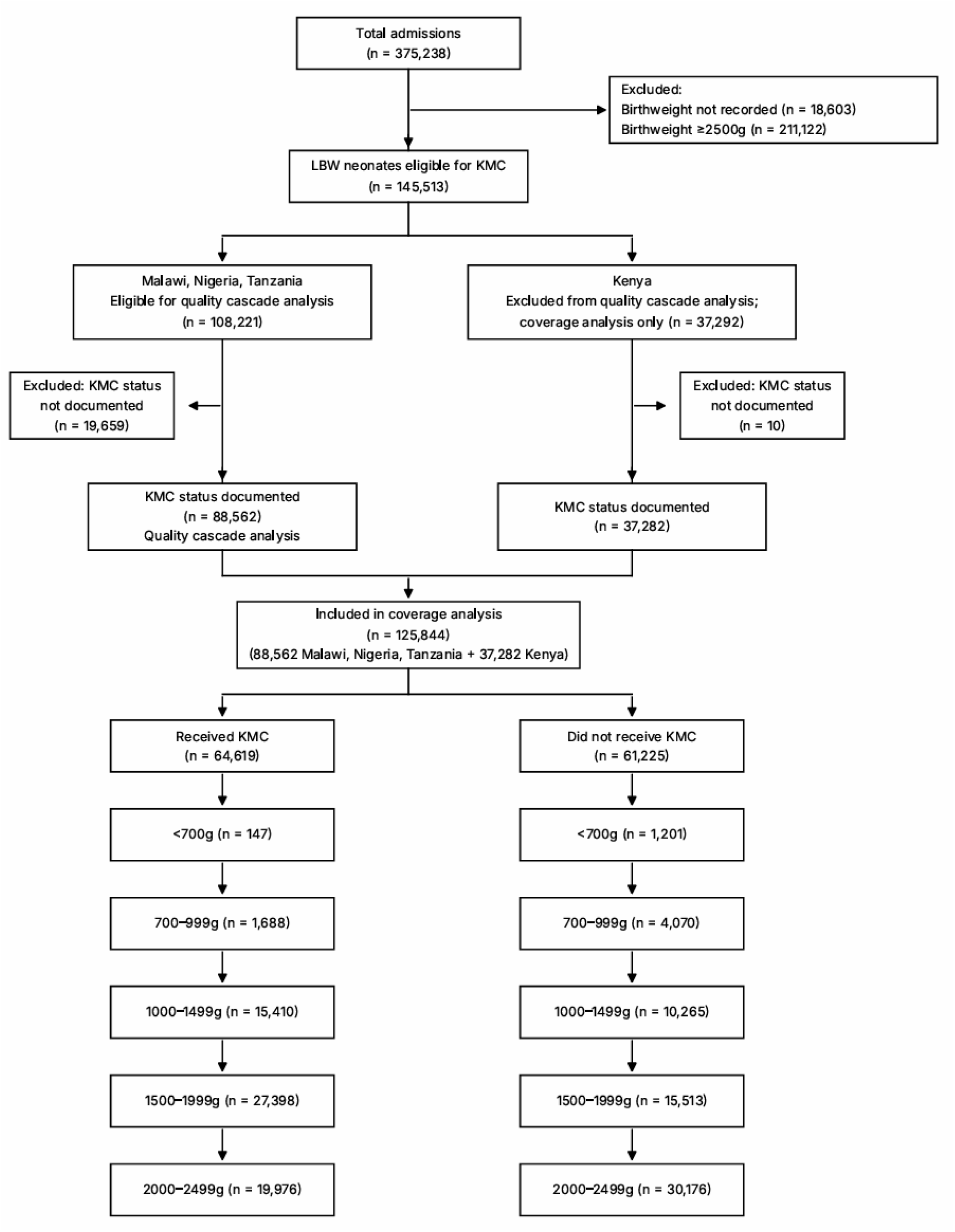
Flow Diagram.

### Results by objectives Objective 1 – KMC coverage

Between 2021 and 2024, KMC coverage increased in all four countries over the study period (Figure 2): from 47.9% to 62.8% in Malawi, 37.2% to 54.5% in Kenya, 33.8% to 61.2% in Tanzania, and 61.1% to 88.7% in Nigeria.

**Figure 2:**
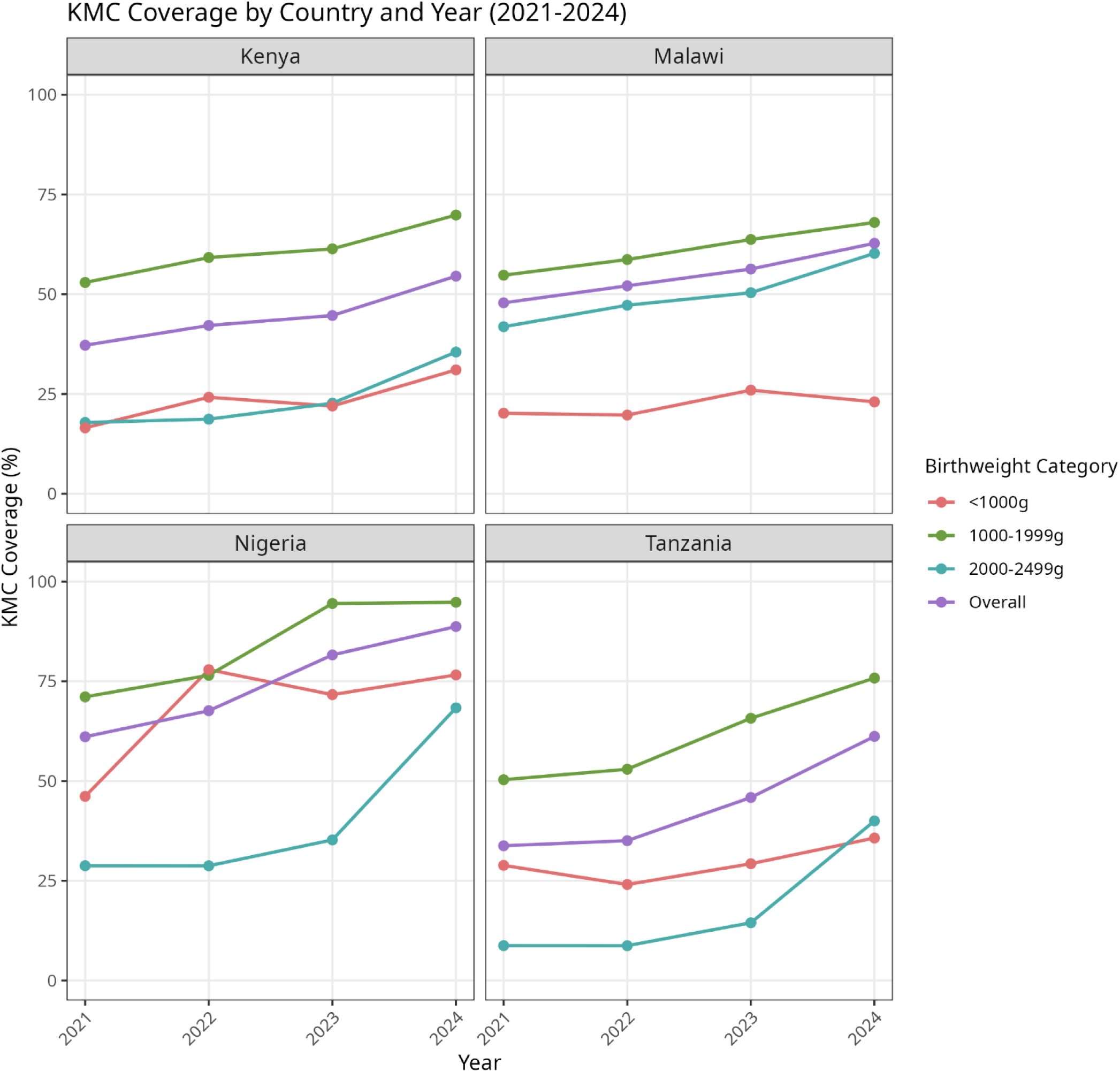
KMC Coverage by Country and Year (2021-2024)

In almost every country and year, coverage was highest among neonates weighing 1000– 1999g, reaching 68.0-94.8% by 2024; the exception was Nigeria in 2022, when coverage was marginally higher among neonates <1000g. Coverage was lower in the <1000g and 2000-2499g categories, though both improved over time. The largest gains were in the 2000-2499g category, particularly in Tanzania (8.8% to 40.0%) and Nigeria (28.8% to 68.3%). Coverage among neonates <1000g remained low in most countries (16.5-35.7%), with the exception of Nigeria (46.2-77.9%).

### Objective 2: Quality KMC Quality Cascade

108,221 neonates were eligible for inclusion in the quality analysis. 88,562 (81.8%) had documentation indicating whether KMC was administered, of whom 48,047 (54.3%) were started on KMC. Among those started on KMC, 44,264 (92.1%) had a valid start date recorded, indicating a relatively small gap in start-date documentation.

Of newborns with a start date, only 12,934 neonates (29.2%) were documented as having KMC initiated on the day of birth. Of these, 8,110 (62.7%) neonates were documented as receiving KMC through to discharge.

Across the entire cascade, only 8,110 (7.5%) of 108,221 KMC-eligible neonates were both initiated on KMC on the first day of life and retained in KMC care until discharge (Figure 3).

**Figure 3:**
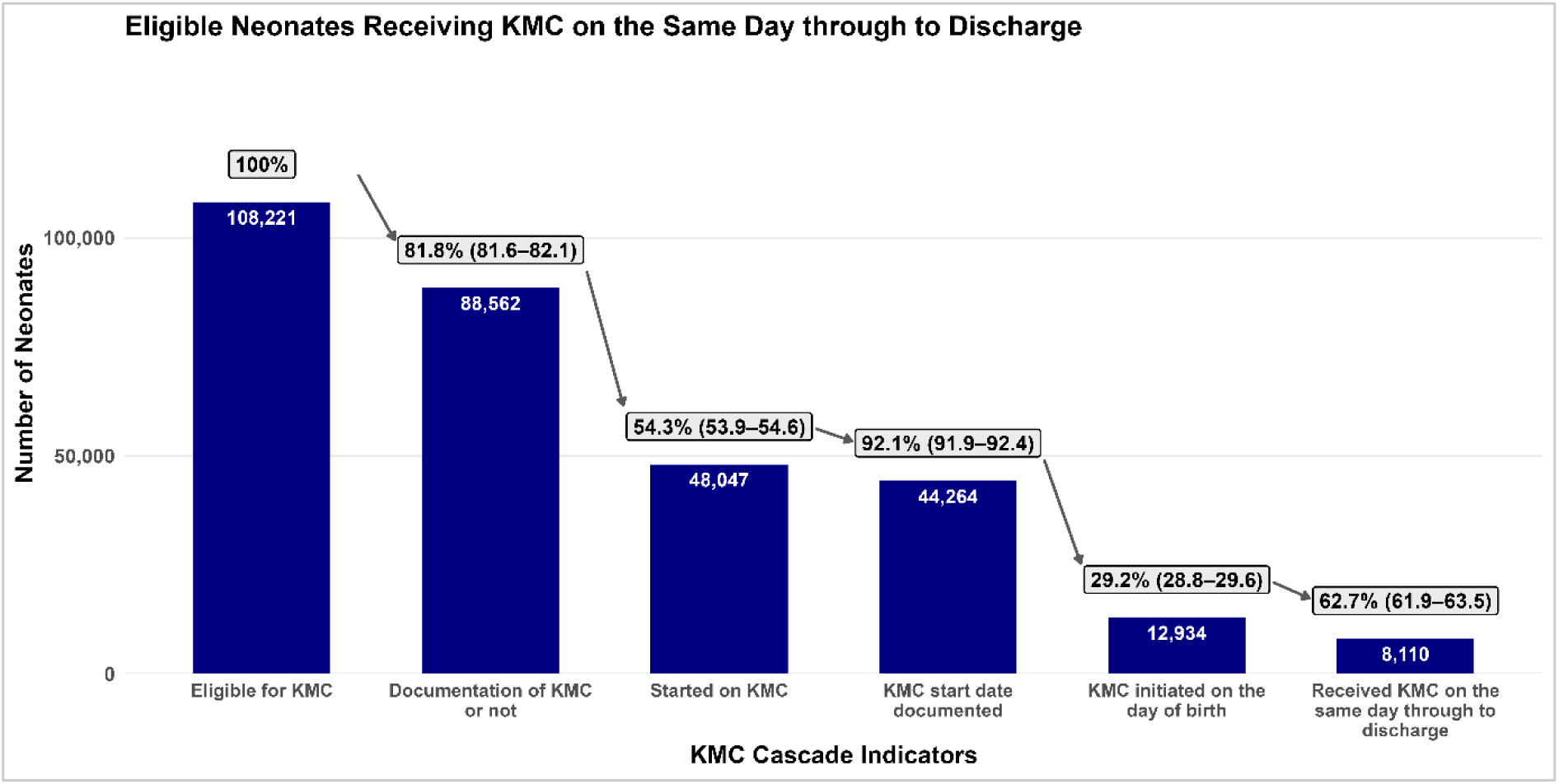
KMC Quality Cascade. Clinical quality cascade reflecting timing of KMC initiation and duration of therapy, pooled across countries (January 2021-December 2024). Initial n= 108,221. In this cascade, the first indicator is the number of KMC-eligible neonates and constitutes 100%. This number then serves as the denominator for the subsequent indicator in the cascade. Each subsequent indicator uses the preceding total as its denominator. Results were pooled across Malawi, Tanzanian, and Nigerian hospitals, with the inter-country range illustrated in parentheses next to overall percentages above bars on the graph.

### KMC and Nutrition Quality Cascade

Among the 44,264 neonates who had a documented KMC start date, 30,003 (67.8%) received breastmilk within the first day of life. Of those who initiated breastmilk early, 29,448 (98.2%) were exclusively breastfed at discharge.

However, because only 44,264 of 108,221 eligible neonates were started on KMC with a documented start date, largely because nearly half of neonates with documented KMC status were not started on KMC, the proportion of all KMC-eligible neonates who both received KMC and were documented as having initiated early breastmilk feeding was substantially lower (Figure 4). Overall, 29,448 (27.2%) of KMC-eligible neonates were initiated on KMC and received breastmilk on the same day, with both interventions continued through to discharge.

**Figure 4:**
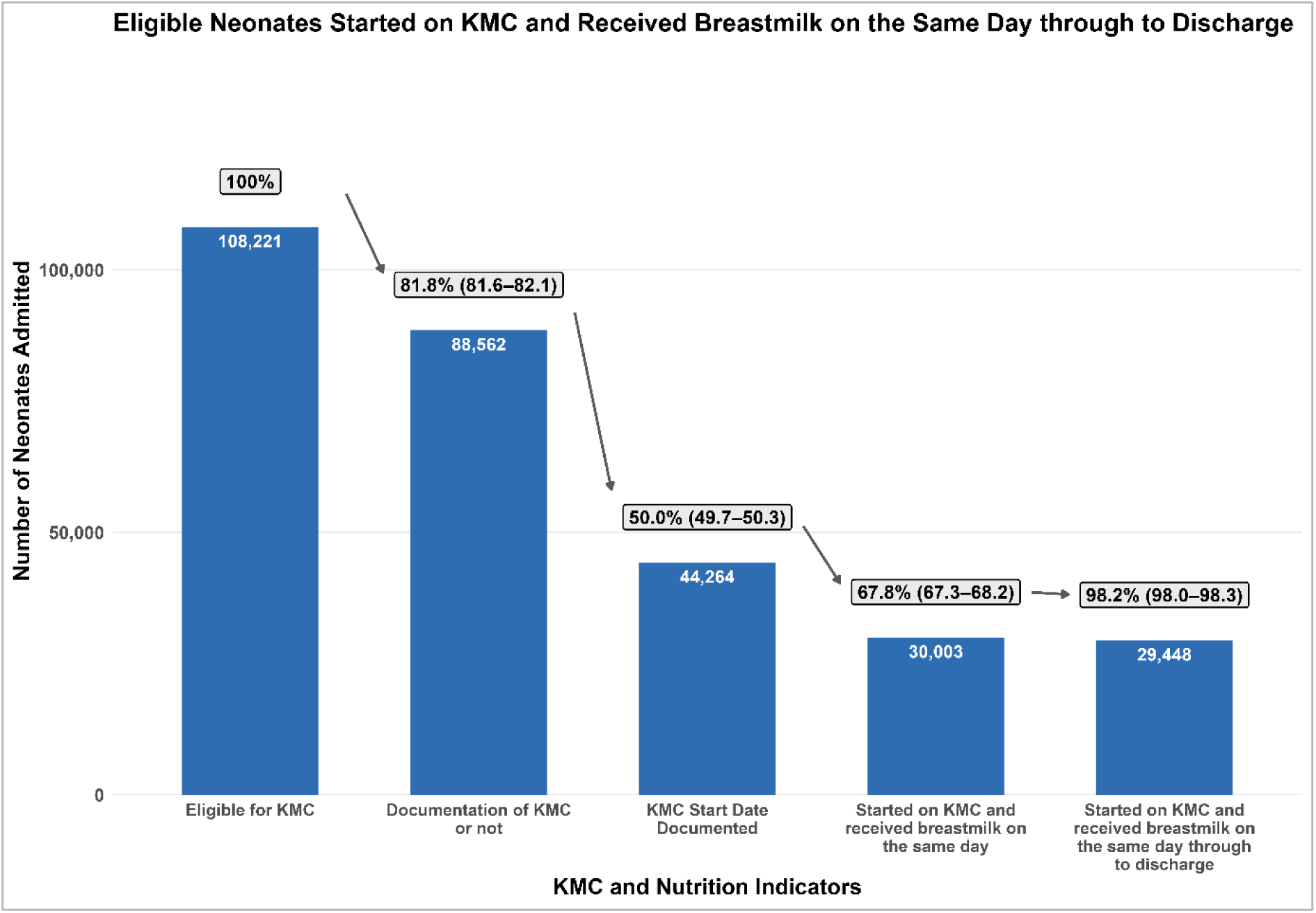
KMC-Nutrition Quality Cascade. Clinical quality cascade reflecting KMC neonates who received exclusive breastmilk on the same day through to discharge, pooled across all countries (January 2021-December 2024). Initial n= 108,221. In this cascade, the first indicator is the number of KMC-eligible neonates and constitutes 100%. This number then serves as the denominator for the subsequent indicator in the cascade. Each subsequent indicator uses the preceding total as its denominator. Results were pooled across Malawi, Tanzanian, and Nigerian hospitals, with the inter-country range illustrated in parentheses next to overall percentages above bars on the graph.

### KMC and CPAP Cascade

Kenyan facilities were excluded from this cascade because KMC timing data were unavailable during the study period. Among the 25,654 neonates eligible for both KMC and CPAP, 22,241 (86.7%) had a documented record of whether or not they received KMC, of whom 11,985 (53.9%) were started on KMC and had start date documented. Of those, only 4,720 (39.4%) also received CPAP during admission, demonstrating that concurrent delivery of KMC and respiratory support was achieved in only a small minority of neonates who required both interventions (Figure 5). Overall, only 4,720 (18.4%) of the 25,654 KMC- and CPAP-eligible neonates had documented receipt of both KMC and CPAP during admission.

**Figure 5:**
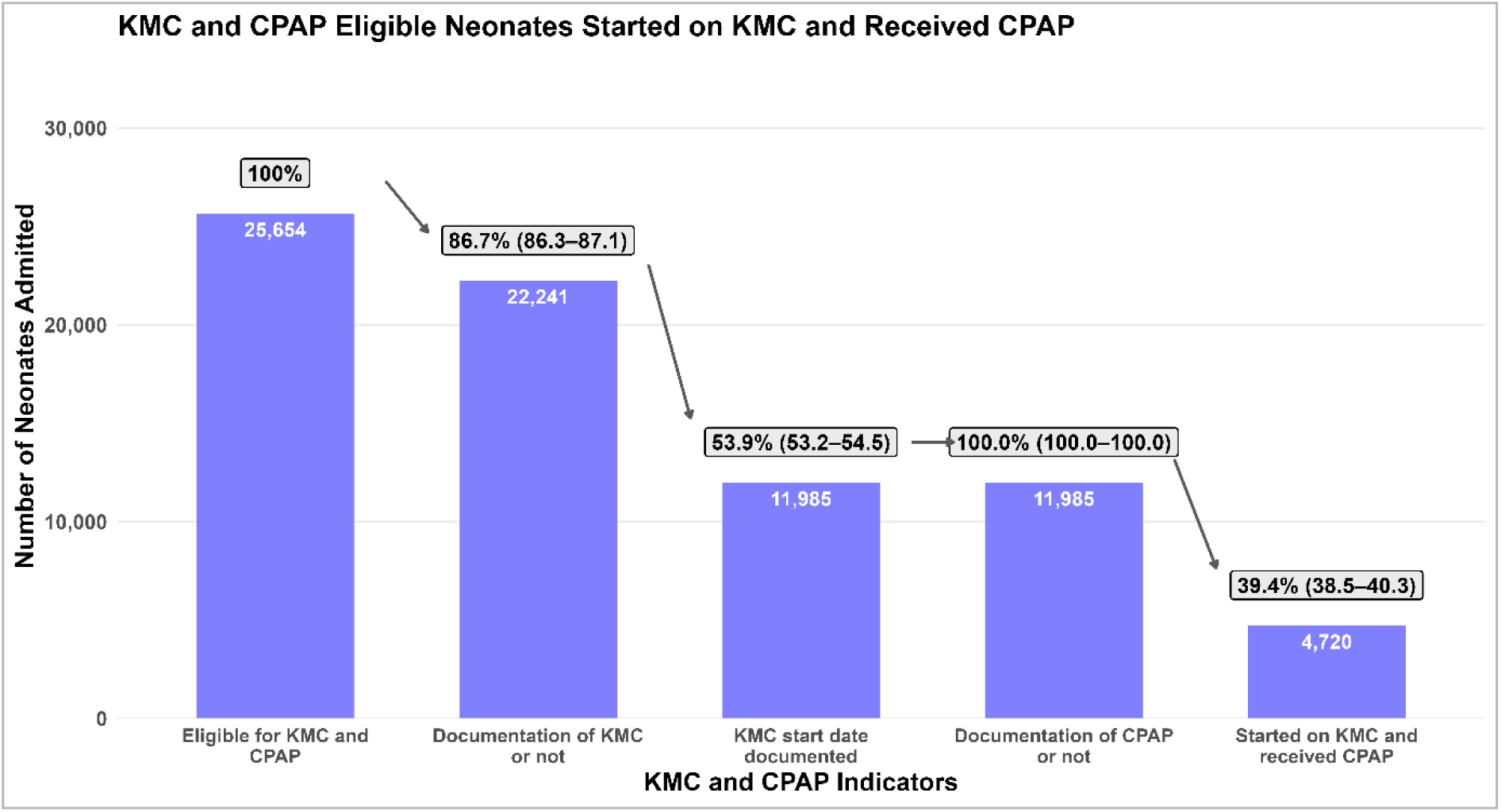
KMC and CPAP Cascade. **Clinical quality cascade reflecting KMC neonates who received CPAP, pooled across all countries (January 2021-December 2024).** Initial n= 25,654. In this cascade, the first indicator is the number of KMC-eligible neonates and constitutes 100%. This number then serves as the denominator for the subsequent indicator in the cascade. Each subsequent indicator uses the preceding total as its denominator. Results were pooled across Malawi, Tanzanian, and Nigerian hospitals, with the inter-country range illustrated in parentheses next to overall percentages above bars on the graph.

### Objective 3: Association between KMC and Survival at Discharge

#### a) Primary Analysis

After adjusting for patient-level characteristics, and clustering within hospitals, neonates documented as receiving KMC consistently demonstrated markedly higher predicted probabilities of survival across all severity categories. Both the absolute and relative survival advantages associated with KMC increased progressively with worsening clinical severity, with the greatest differences observed among severely ill neonates (Table 1).

**Table 1:** Adjusted predicted probability of survival, % (95% CI)

|  | **Mild Severity (3–4)** | **Moderate Severity (5–6)** | **High Severity (7–9)** |
|---|---|---|---|
| **Documented no KMC** | 77.6% (75.3–79.7) | 30.0% (27.4–32.8) | 7.0% (5.8–8.4) |
| **KMC received** | 97.3% (96.9–97.6) | 91.1% (89.9–92.1) | 79.5% (75.8–82.7) |
| **Absolute Difference** | +19.7% | +61.1% | +72.5% |

**Note:** Values are adjusted predicted probabilities of survival at discharge with 95% confidence intervals, estimated from a mixed-effects logistic regression model including KMC receipt, baseline illness severity, sex, birth location, and the interaction between KMC receipt and illness severity. Facility was included as a random intercept to account for clustering of neonates within hospitals. “Documented no KMC” refers to neonates for whom KMC administration was documented as not received; neonates with missing KMC administration status were excluded from this analysis

Overall, after adjustment for patient-level characteristics, and hospital-level clustering, KMC receipt was associated with higher survival across all illness severity groups (Tables 1 and 2). The association between KMC and survival varied significantly by baseline illness severity, with the magnitude of the association increasing with greater severity. Among neonates with mild illness, KMC was associated with substantially higher odds of survival (aOR 10.39, 95% CI 9.67–11.16), with significant interaction effects observed for moderate (interaction aOR 2.29, 95% CI 2.06–2.55) and severe illness (interaction aOR 4.99, 95% CI 3.91–6.35). Increasing illness severity was independently associated with lower odds of survival. Male and outborn neonates also had lower adjusted odds of survival than female and inborn neonates, respectively (Table 2).

**Table 2:** Adjusted Associations between KMC and survival at discharge.

| **Model term** | **Adjusted Odds Ratio (aOR)** | **95% CI** | **p-value** |
|---|---|---|---|
| KMC received vs documented no KMC at mild severity | 10.39 | (9.67, 11.16) | <0.001 |
| KMC × moderate severity interaction | 2.29 | (2.06, 2.55) | <0.001 |
| KMC × severe severity interaction | 4.99 | (3.91, 6.35) | <0.001 |
| Moderate severity vs mild severity, among neonates with documented no KMC | 0.124 | (0.118, 0.131) | <0.001 |
| Severe severity vs mild severity, among neonates with documented no KMC | 0.022 | (0.018, 0.025) | <0.001 |
| Male vs Female | 0.89 | (0.85, 0.92) | <0.001 |
| Outborn vs Inborn | 0.53 | (0.51, 0.56) | <0.001 |

**Note:** Adjusted odds ratios were estimated using a mixed-effects logistic regression model with survival at discharge as the binary outcome. The model included KMC receipt, baseline illness severity, the interaction between KMC receipt and illness severity, sex, and birth location as fixed effects, with hospital included as a random intercept to account for clustering of neonates within hospitals. Reference categories were documented no KMC, mild severity, female, and inborn. Baseline illness severity was derived from a composite score incorporating gestational age, oxygen saturation at admission, and admission temperature, and categorized as mild (3–4), moderate (5–6), or severe (7–9). The KMC-by-severity interaction significantly improved model fit compared with the model without the interaction (Likelihood Ratio Test χ² = 338.84, df = 2, p < 0.001). KMC = kangaroo mother care; aOR = adjusted odds ratio; CI = confidence interval.

##### Predicted Probability of Survival

Figure 6 presents the adjusted predicted probability of survival across neonatal severity groups for babies who received KMC versus those who did not receive KMC, after adjustment for illness severity, their interaction, sex, and birth location, with hospital included as a random intercept.

**Figure 6:**
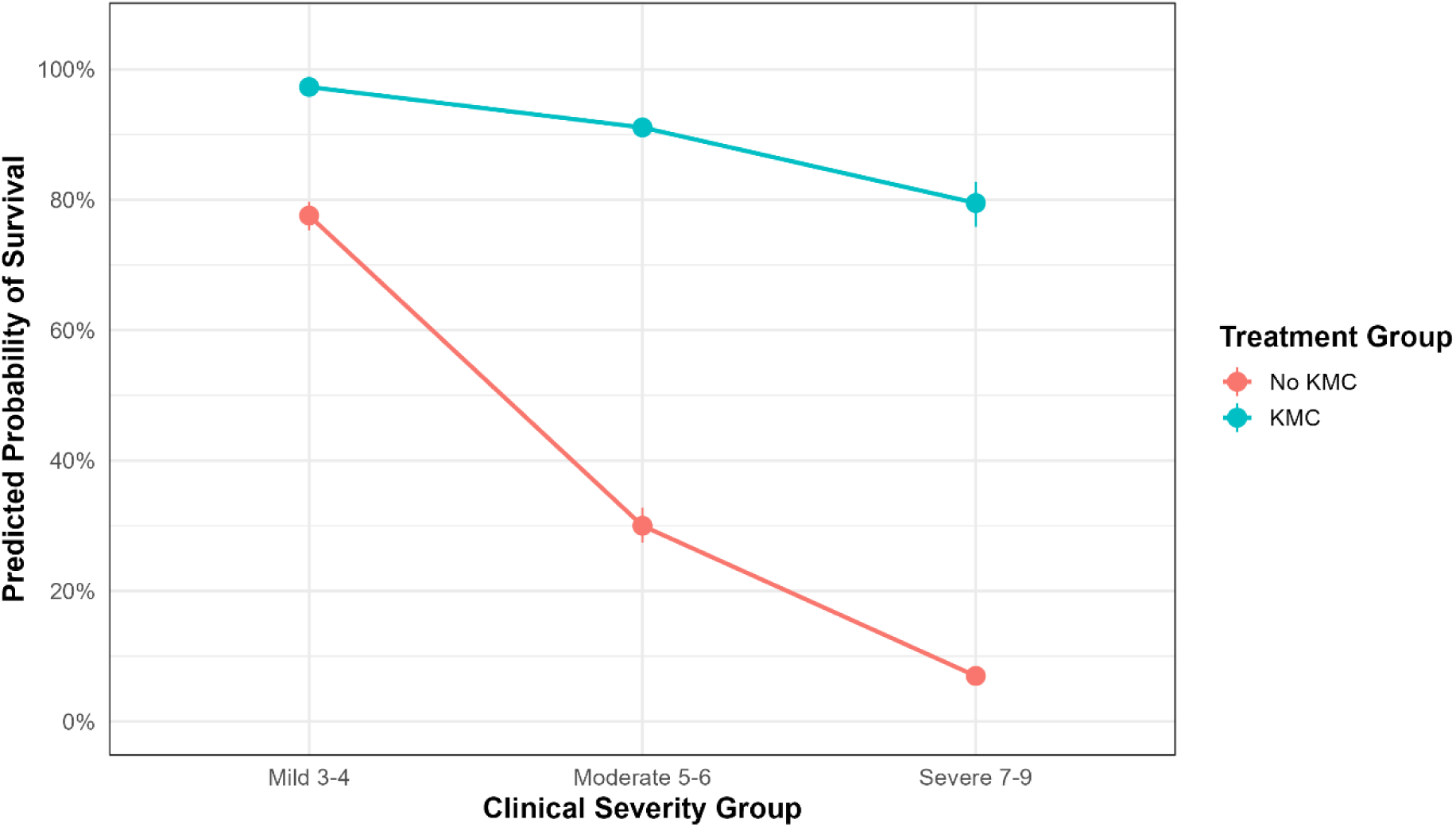
Adjusted Predicted Survival Probability of KMC, Severity, Patient--levels Covariates.

Adjusted predicted probabilities from the mixed-effects logistic regression model demonstrated progressively lower survival with increasing clinical severity among neonates who did not receive KMC. In contrast, neonates receiving KMC maintained substantially higher predicted survival across all severity groups. The magnitude of the difference between KMC and non-KMC groups increased with severity, with the largest separation observed among severely ill neonates, supporting a significant interaction between KMC and severity category. The gap between the KMC and non-KMC groups widens as severity increases. The association between KMC and survival becomes stronger with increasing neonatal severity.

##### The ROC (Receiver Operating Characteristics) Analysis

The mixed-effects KMC survival model demonstrated good discrimination, with an area under the ROC curve (AUC) of 0.8699, indicating that the model has an 87% probability of correctly distinguishing between a neonate who survived and one who did not survive (Appendix 3).

#### b) Secondary Analysis

##### Time of KMC Initiation

Among neonates who received KMC, later KMC initiation was associated with higher adjusted survival compared with same-day initiation after accounting for baseline illness severity, birthweight, sex, birth location, and facility-level clustering. Adjusted predicted probabilities showed lower survival with increasing baseline illness severity across KMC initiation groups, with the lowest predicted survival observed among severely ill neonates receiving same-day KMC (Figure 7).

**Figure 7:**
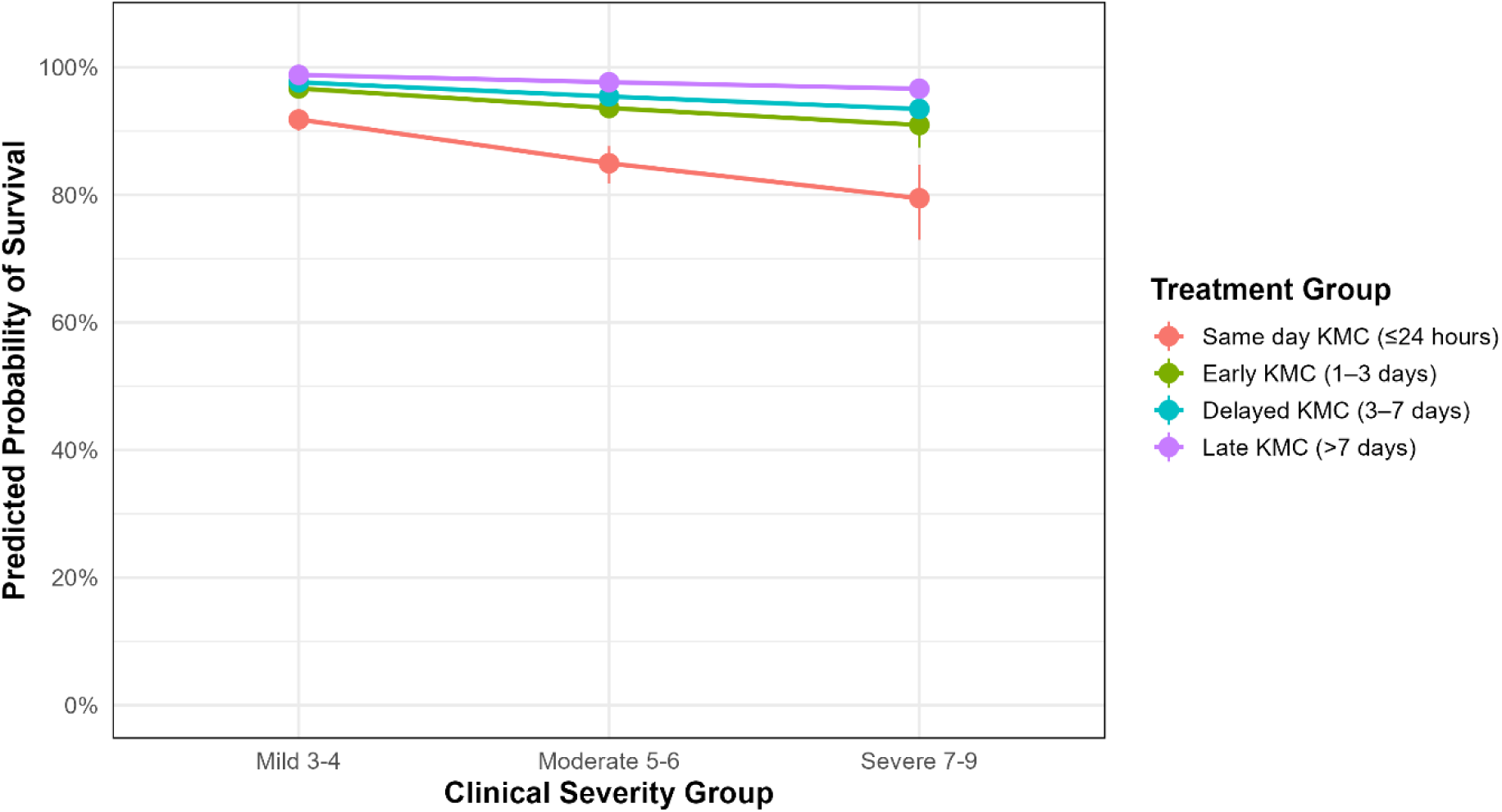
Adjusted Predicted Survival Probability by Severity and Time to KMC Initiation.

However, the apparent survival advantage associated with later initiation should be interpreted cautiously, as neonates in the delayed and late initiation groups had necessarily survived long enough to receive KMC, introducing potential survivor selection bias. Detailed adjusted estimates are presented in Appendix 4. In these analyses, later KMC initiation remained positively associated with survival, while greater illness severity, male sex, and outborn status were associated with lower survival. Birthweight was also strongly associated with survival, with progressively higher adjusted odds among higher birthweight categories (Appendix 4). Birthweight-stratified predicted probabilities further showed that differences in survival by timing of KMC initiation were most pronounced among smaller and more severely ill neonates (Appendix 4). These secondary findings should therefore be interpreted as associations rather than evidence that delaying KMC improves survival. To support the use of these findings in clinical training and advocacy, a plain-language summary of the key associations has been developed and is presented in Appendix 5.

## DISCUSSION

This study presents a large multi-country analysis of KMC coverage and quality in sub-Saharan Africa, drawing on prospectively collected, individually-linked data from 66 hospitals across four countries over a four-year period. By adapting quality-of-care cascades such as used from HIV, to KMC and to other neonatal care interventions [31–33] , this analysis quantifies where, and to what extent, eligible neonates are lost from the care pathway. These results challenge the tendency to treat KMC coverage as a unitary indicator and demonstrates the analytical value of disaggregating the care pathway to identify where, and for whom, delivery of evidence-based KMC care consistently falls short.

Across these 65 sites in the four countries, KMC coverage increased over the study period, with Nigeria and Tanzania recording the largest absolute increases (27.6 and 27.4 percentage points, respectively). Malawi’s higher baseline coverage, with steady and sustained improvement in coverage builds on long-standing national institutionalisation of KMC, having integrated the intervention into national guidelines as early as 2005, and pre-service training established. This extended implementation history may have fostered greater acceptability among both healthcare staff and caregivers, and created conditions in which localised adaptations, such as hospital-level innovations in KMC positioning support, could be established and sustained [34–36]. Nigeria’s coverage figures may reflect national newborn care policies and the establishment of KMC champions at hospital level [37,38]. Tanzania’s improvements in overall coverage, alongside coverage in the 2000–2499g category that, despite marked improvement, remained below that of other weight categories, illustrate a pattern observed across settings in which policy revision precedes changes in clinical practice, as the 2022 extension of KMC eligibility to all infants under 2500g does not appear to have been fully operationalised in this weight category within the study period. In Kenya, coverage increased gradually but at a slower pace than other countries, reflecting documented challenges in translating national policy into consistent frontline practice, including resource constraints, limited staff time, which have been identified as barriers to sustained uptake [39]. Infrastructural limitations have also been identified as barrier to high quality KMC in Kenya [40]. Notably, Kenya’s expansion of free maternity services has increased demand for hospital-based newborn care, and it is plausible that growth in the volume of eligible neonates outpaced the expansion of KMC capacity, contributing to lower overall coverage rates [41]. It is important to note that these findings are drawn entirely from NEST data collection, which operated at scale only in Malawi; coverage trends for Kenya, Nigeria, and Tanzania therefore may not be representative of national-level trends in those countries. Taken together, these country-level findings underscore that coverage trajectories are shaped by the intersection of policy maturity, health system capacity, and equity, and that strategies to improve KMC uptake must be context-specific.

The quality cascade findings carry direct implications for how KMC programmes in LMICs are designed, monitored, and resourced. The most striking observation is the marked gap in the timeliness of initiation: only around one in eight eligible neonates (12.0%) were recorded to be initiated on KMC on the first day of life, and fewer than one in ten (7.5%) were initiated on the first day and continued through to discharge, despite guidance explicitly recommending initiation as soon as possible after birth [11]. The finding that most KMC in this cohort was not initiated on the first day of life may not be due to clinical prioritisation, but the practical consequences of an infrastructure gap. The 2022 WHO policy recommending KMC prior to clinical stability was published only shortly before the latter years of this study’s analysis period, and evidence from comparable level-2 care settings in sub-Saharan Africa suggests that substantial hospital-level infrastructure improvements are required to implement KMC prior to stability safely [8,42]. Where the conditions for safe early initiation do not yet exist, premature implementation carries clinical risk, and programmes must therefore ensure that advocacy for earlier KMC is accompanied by proportionate investment in the enabling and necessary infrastructure. Programmes should also consider incorporating timeliness of initiation as a standard quality indicator alongside coverage, shifting monitoring frameworks towards the concept of effective coverage, which accounts for both coverage and the quality of the care delivered.

Infrastructure and workforce readiness are critical determinants of whether KMC can be delivered consistently at the hospital level. Service readiness assessments across these same hospitals and time periods have highlighted substantial gaps in the infrastructure needed to support small and sick newborn care [31]. Bed occupancy was highly variable, with almost a third of hospital-months exceeding full capacity, and nursing staffing ratios, particularly at night, frequently fell short of recommended levels, especially in Kenya [31]. A comprehensive Health Facility Assessment (HFA) tool for small and sick newborn care, developed and operationalised by NEST360, has also been used to systematically evaluate hospital readiness across the same 66 hospitals in Kenya, Malawi, Nigeria, and Tanzania [43]. At baseline, overall service readiness for small and sick newborn care was a median of 51% [IQR 48–57%] [19]. Family-centred care, which encompasses KMC, parental involvement, and the support systems needed to sustain these practices, was the most poorly resourced component of neonatal care across all four countries at just 27% [IQR 18–40%] [19].These gaps extended beyond equipment to the workforce capacity and physical environment needed to support mothers and families in practising KMC, highlighting that hospital readiness for KMC remains substantially limited across these settings. Translating policy into consistent practice requires a paradigm shift towards care of the mother-infant dyad, supported by a whole-systems approach addressing workforce, infrastructure, and institutional culture.

The co-intervention cascades provide additional insight into the co-coverage of newborn care interventions. Within the nutrition cascade, 67.8% of neonates with a documented KMC start date received breastmilk on the first day of life, and of those, 98.2% were exclusively breastfed at discharge. While these figures are encouraging, they must be interpreted in the context of the substantially reduced denominator at this stage of the cascade. The apparent success in breastfeeding applies only to those neonates who had already progressed through earlier documentation and initiation steps, and therefore could reflect a selected group receiving more consistent overall care. Early initiation of breastfeeding within the first hour of life is associated with sustained exclusive breastfeeding and reduced neonatal mortality [44,45]. However, NEST360 currently records breastmilk initiation only by day of life, meaning that this one-hour target cannot be assessed. Capturing timing of first feed in hours would enable hospitals to determine whether practice aligns with this recommendation and to identify modifiable delays in early initiation. Within the CPAP cascade, among neonates receiving KMC who were also assessed as requiring CPAP, about 39.4% had CPAP administered. As with the primary cascade, it is not possible to determine whether this reflects under-delivery of respiratory support, incomplete documentation, or both. However, given that a significant proportion of KMC-eligible neonates will also require concurrent respiratory support, this figure warrants attention and reinforces the case for strengthening both clinical capacity and documentation systems for co-interventions alongside KMC.

The greatest loss along the cascade occurred at KMC initiation, where 45.7% of neonates with documented KMC status were not started on KMC, considerably larger than the loss attributable to documentation gaps (18.2%). This indicates that non-provision of KMC, rather than failure to record it, is the major driver of the coverage shortfall observed. Nonetheless, documentation gaps remain a systemic challenge in routine data capture rather than a problem specific to any single intervention, and routine data cannot fully distinguish non-delivery of care from failure to document care that was provided. Evidence from comparable settings indicates that the absence of standardised recording tools and reporting processes is a recognised barrier to monitoring progress at scale [46,47]. Strengthening and standardising documentation of KMC must therefore be treated as an integral component of programme design and scale up [48–50], both to improve the accuracy of coverage estimates and to ensure that monitoring frameworks can meaningfully distinguish between failure to deliver and failure to record.

Survival outcomes were analysed across birthweight and severity strata. These findings suggest that the observed relationship between KMC timing and survival is strongly influenced by baseline neonatal risk, with timing becoming less prognostically important among larger neonates who generally experienced favourable survival outcomes across all timing groups. These findings also suggest that although increasing illness severity markedly reduced neonatal survival, KMC remained strongly associated with improved survival and appeared to provide the greatest relative benefit among the most critically ill neonates.

The findings of this study must also be considered in light of recent evidence on the benefits of KMC prior to stability. Recent evidence indicates that immediate KMC for infants weighing 1.0 to 1.799kg, regardless of clinical stability, may reduce neonatal mortality by up to 25% [28]. While KMC prior to stabilisation does not significantly affect early neonatal mortality at fewer than seven days, likley since these very vulnerable babies need more intensive care, it has been shown to reduce in 28-day mortality in both iKMC trial and OMWANA, especially with higher duration [8]. These findings strengthen the case for earlier initiation and higher duration each day during admission and after lend urgency to the infrastructure investments discussed above. The fact that fewer than one in ten eligible neonates in this cohort received KMC on the first day of life must therefore be understood not only as a quality gap, but as a missed opportunity to deliver an intervention with meaningful survival benefit.

Our analysis has a number of methodological strengths. The scale and longitudinal nature of the dataset, encompassing over 145,000 KMC-eligible neonates across 66 hospitals and four years, provides statistical power and cross-country comparability. Unlike aggregated national estimates, individually linked inpatient data from the NID allow each neonate to be followed from admission to discharge or death, enabling multi-stage quality cascades, adjustment for severity, and identification of the specific points at which care is lost. This granularity represents the principal contribution of the NID to understanding, and improving, the delivery of small and sick newborn care. The development of coverage indicators for KMC within routine health information systems remains an active area of international policy work, with individual case-note data recognised as the most appropriate source for capturing inpatient KMC indicators, despite known data quality challenges [46].

This study demonstrates that structured improvement programmes can achieve meaningful and sustained increases in KMC coverage across diverse health system contexts in sub-Saharan Africa. However, the cascade analyses presented here reveal that coverage, as conventionally measured, is an insufficient indicator of programmatic success. The stepwise losses from eligibility to documented, timely, and continuous KMC delivery illustrate that the distance between a policy commitment to KMC and its realisation as effective care remains considerable. The 2023 WHO Global Position Paper on KMC calls for a fundamental reorganisation of maternal-newborn care, with immediate initiation following birth as the new standard of practice [51]. Achieving this standard across high-burden settings will require investment not only in clinical training and commodities, but in the data systems, institutional accountability structures, and enabling environments that include infrastructural changes to support the mother-infant dyad.

Several limitations must be acknowledged. Firstly, there was a substantial amount of missing data for KMC and birthweight. It is plausible that the most critically ill and smallest neonates were least likely to have accurately recorded birthweight and KMC data, which would mean these neonates are under-represented in the analysed sample and could bias the birthweight- and severity-stratified findings. KMC timing data were unavailable for Kenyan facilities, which were therefore excluded from all cascade analyses. Elsewhere, start dates were documented for 92.1% of neonates started on KMC, so missing timing data are unlikely to have substantially affected estimates of timely initiation, although the proportion initiated on the first day of life (12.0%) should be considered as underestimated. Data completeness in routine hospital records across LMICs is highly heterogeneous, and incomplete recording in non-standardised case notes may systematically underrepresent indicator values [52]. These limitations highlight gaps in the infrastructure that support consistent care delivery. The analysis is further limited to NEST360 phase 1 implementation hospitals, which received structured programme support to support KMC uptake and are therefore unlikely to be representative of the broader hospital landscape in each country. Findings should be interpreted in this context and not generalised to non-programme settings without caution.

A limitation is the absence of a KMC duration indicator generally and also within NEST360 data systems. Duration of skin-to-skin contact per day is a major determinant of KMC health impact [53,54], which reflects a current gap in NEST360 data collection. To accurately capture KMC duration in future, practical approaches such as integrating duration recording into routine documentation should be considered, a practice that has proven reasonably accurate in a validation study in Uganda [55,56]. Novel digital tools could monitor skin-to skin contact and hence measure KMC duration more reliably [57]. NEST360 plans to test KMC duration measures and include in the NID, and importantly use the data to increase KMC impact.

For the world’s smallest and most vulnerable newborns, the gap between what is known to work and what is delivered results in lives lost. Closing that gap demands a shift from measuring whether KMC is offered to ensuring coverage and quality are tracked and data used for KMC for every eligible newborn at the right time and duration in every hospital, and country. As national targets to ending preventable neonatal deaths approach their 2030 deadline, the evidence presented here underscores that scale-up of small and sick newborn care must be accompanied by sustained investment in quality, accountability, and the health systems capable of delivering both.

For the world’s smallest and most vulnerable newborns, the gap between what is known to work and what is consistently delivered represents a preventable loss of life. Closing that gap demands a shift from measuring whether KMC is offered to ensuring it is delivered equitably, safely, and at the right time, for every eligible newborn in every country. As global commitments to ending preventable neonatal deaths approach their 2030 deadline, the evidence presented here underscores that scale-up of small and sick newborn care must be accompanied by sustained investment in quality, accountability, and the health systems capable of delivering both.

## Data Availability

All data produced in the present study are available upon reasonable request to the authors

## LIST OF ABBREVIATIONS

aOR: Adjusted odds ratio
CI: Confidence interval
CPAP: Continuous positive airway pressure ENAP *Every Newborn* Action Plan
EWENE: Every Woman, Every Newborn, Everywhere KMC Kangaroo Mother Care
LBW: Low birthweight
LMICs: Low and middle income countries
NEST360: Newborn Essential Solutions and Technologies NID Neonatal Inpatient Dataset
OR: Odds ratio
SDGs: Sustainable Development Goals WHO World Health Organization

## DECLARATIONS

### Ethics and consent to participate

NEST360 complex was granted ethical approval from the Institutional Review Boards of all four countries, the London School of Hygiene & Tropical Medicine and Rice University.

All collaborators received local ethical permissions for their data (Appendix 6).

## Consent for publication

Not applicable.

## Availability of data and material

Data sharing and transfer agreements were jointly developed and signed by all collaborating partners. The data may be available upon request from LSHTM data compass systems pending approval by a multi-country NEST360 Data Sharing and Publications Group. Data Tools are open access on www.NEST360.org and www.newborntoolkit.org. For the purpose of open access, the author has applied a ‘Creative Commons Attribution’ (CC BY) licence to any Author Accepted Manuscript version arising.

## Authors’ contributions

This analysis was done as part of NEST360 Alliance and partners. ER, JT, SC, and CAB developed the research questions and overall analysis plan for this paper, with KS and JEL as part of neonatal intervention coverage and quality cascades work and severity score used in four papers in this BMC supplement. Analysis was led by ER and JT. ER, JT, and CAB drafted the manuscript. JEL and NR provided valuable guidance and support on the analyses and manuscript development. All authors reviewed and helped to revise the manuscript. All authors reviewed and agreed on the final version. The authors’ views are their own, and not necessarily from any of the institutions they represent.

## Acknowledgements

Firstly, and most importantly, we thank the women, families, and newborns together with all the health workers involved in this work. Finally, we are grateful to fellow researchers and guest editors who reviewed this paper, and for the input from managing editors at BMC and within NEST360 including Caroline Noxon, Kristina Shemwell, Harriet Ruysen, William Macharia, and Joy E. Lawn.

## APPENDIX

Appendix 1: STROBE Checklist

| **Section** | **Item No** | **Recommendation** | **Page No** |
|---|---|---|---|
| Title and abstract | 1 | (a) Indicate the study's design with a commonly used term in the title or the abstract<br>(b) Provide in the abstract an informative and balanced summary of what was done and what was found | 1-2<br>1-3 |
| **Introduction** |  |  |  |
| Background/rationale | 2 | Explain the scientific background and rationale for the investigation being reported | 5-7 |
| Objectives | 3 | State specific objectives, including any prespecified hypotheses | 7-8 |
| **Methods** |  |  |  |
| Study design | 4 | Present key elements of study design early in the paper | 8-12 |
| Setting | 5 | Describe the setting, locations, and relevant dates, including periods of recruitment, exposure, follow-up, and data collection | 8-12 |
| Participants | 6 | (a) Cohort study—Give the eligibility criteria, and the sources and methods of selection of participants. Describe methods of follow-up<br>Case-control study—Give the eligibility criteria, and the sources and methods of case ascertainment and control selection. Give the rationale for the choice of cases and controls<br>Cross-sectional study—Give the eligibility criteria, and the sources and methods of selection of participants<br>(b) Cohort study—For matched studies, give matching criteria and number of exposed and unexposed<br>Case-control study—For matched studies, give matching criteria and the number of controls per case | 12-13 |
| Variables | 7 | Clearly define all outcomes, exposures, predictors, potential confounders, and effect modifiers. Give diagnostic criteria, if applicable | 8-13 |
| Data sources/measurement | 8* | For each variable of interest, give sources of data and details of methods of assessment (measurement). Describe comparability of assessment methods if there is more than one group | 8-13 |
| Bias | 9 | Describe any efforts to address potential sources of bias | 10 |
| Study size | 10 | Explain how the study size was arrived at | 12, Figure 1 |
| Quantitative variables | 11 | Explain how quantitative variables were handled in the analyses. If applicable, describe which groupings were chosen and why | 9-12 |
| Statistical methods | 12 | (a) Describe all statistical methods, including those used to control for confounding<br>(b) Describe any methods used to examine subgroups and interactions<br>(c) Explain how missing data were addressed<br>(d) Cohort study—If applicable, explain how loss to follow-up was addressed<br>Case-control study—If applicable, explain how matching of cases and controls was addressed<br>Cross-sectional study—If applicable, describe analytical methods taking account of sampling strategy<br>(e) Describe any sensitivity analyses | 10-12<br>11-12<br>12<br>8-12 |
| **Results** |  |  |  |
| Participants | 13* | (a) Report numbers of individuals at each stage of study—eg numbers potentially eligible, examined for eligibility, confirmed eligible, included in the study, completing follow-up, and analysed<br>(b) Give reasons for non-participation at each stage<br>(c) Consider use of a flow diagram | 11-12<br>Figure 1<br>Figure 1 |
| Descriptive data | 14* | (a) Give characteristics of study participants (eg demographic, clinical, social) and information on exposures and potential confounders<br>(b) Indicate number of participants with missing data for each variable of interest<br>(c) Cohort study—Summarise follow-up time (eg, average and total amount) | Appendix 2<br>Appendix 2 |
| Outcome data | 15* | Cohort study—Report numbers of outcome events or summary measures over time. Case-control study—Report numbers in each exposure category, or summary measures of exposure. Cross-sectional study—Report numbers of outcome events or summary measures | 14-23 |
| Main results | 16 | (a) Give unadjusted estimates and, if applicable, confounder-adjusted estimates and their precision (eg, 95% confidence interval). Make clear which confounders were adjusted for and why they were included<br>(b) Report category boundaries when continuous variables were categorized<br>(c) If relevant, consider translating estimates of relative risk into absolute risk for a meaningful time period | 12-23<br>n/a<br>n/a |
| Other analyses | 17 | Report other analyses done—eg analyses of subgroups and interactions, and sensitivity analyses | n/a |
| **Discussion** |  |  |  |
| Key results | 18 | Summarise key results with reference to study objectives | 23-31 |
| Limitations | 19 | Discuss limitations of the study, taking into account sources of potential bias or imprecision. Discuss both direction and magnitude of any potential bias | 28-29 |
| Interpretation | 20 | Give a cautious overall interpretation of results considering objectives, limitations, multiplicity of analyses, results from similar studies, and other relevant evidence | 28-29 |
| Generalisability | 21 | Discuss the generalisability (external validity) of the study results | 28 |
| **Other information** |  |  |  |
| Funding | 22 | Give the source of funding and the role of the funders for the present study and, if applicable, for the original study on which the present article is based | 31 |

Appendix 2: Characteristics of Admitted Neonates (January 2021-December 2024)

| Characteristic | Overall<br>N = 125,844<sup>1</sup> | Malawi<br>N = 68,274<sup>1</sup> | Kenya<br>N = 37,282<sup>1</sup> | Tanzania<br>N = 17,613<sup>1</sup> | Nigeria<br>N = 2,675<sup>1</sup> |
|---|---|---|---|---|---|
| **Sex** |  |  |  |  |  |
| Male | 60,154 (49%) | 31,567 (49%) | 18,790 (50%) | 8,496 (48%) | 1,301 (49%) |
| Female | 62,115 (51%) | 33,238 (51%) | 18,418 (49%) | 9,087 (52%) | 1,372 (51%) |
| Indeterminate | 57 (<0.1%) | 23 (<0.1%) | 14 (<0.1%) | 19 (0.1%) | 1 (<0.1%) |
| Unknown | 3,518 | 3,446 | 60 | 11 | 1 |
| **Birthweight category** |  |  |  |  |  |
| <1000g | 7,106 (5.6%) | 2,640 (3.9%) | 2,938 (7.9%) | 1,289 (7.3%) | 239 (8.9%) |
| 1000–1499g | 25,675 (20%) | 12,775 (19%) | 7,904 (21%) | 4,039 (23%) | 957 (36%) |
| 1500–1999g | 42,911 (34%) | 22,512 (33%) | 13,138 (35%) | 6,276 (36%) | 985 (37%) |
| 2000–2499g | 50,152 (40%) | 30,347 (44%) | 13,302 (36%) | 6,009 (34%) | 494 (18%) |
| **Birthweight (g), mean (SD)** | **1,762 (454)** | **1,806 (437)** | **1,724 (473)** | **1,704 (458)** | **1,538 (422)** |
| **Gestational age category** |  |  |  |  |  |
| Extremely preterm (<28 weeks) | 7,233 (6.5%) | 2,751 (4.8%) | 3,351 (9.6%) | 940 (5.7%) | 191 (7.6%) |
| Very preterm (28 to <32 weeks) | 22,743 (21%) | 9,910 (17%) | 8,156 (23%) | 3,692 (23%) | 985 (39%) |
| Late preterm (32 to <37 weeks) | 53,508 (48%) | 26,637 (47%) | 16,822 (48%) | 8,903 (54%) | 1,146 (45%) |
| Term (≥37 weeks) | 27,249 (25%) | 17,603 (31%) | 6,594 (19%) | 2,846 (17%) | 206 (8.1%) |
| Unknown | 15,111 | 11,373 | 2,359 | 1,232 | 147 |
| **Gestational age (weeks), mean (SD)** | **33.7 (16.8)** | **34.3 (19.7)** | **32.9 (4.1)** | **33.6 (22.9)** | **31.8 (3.3)** |
| Unknown | 15,111 | 11,373 | 2,359 | 1,232 | 147 |
| **Age at admission** |  |  |  |  |  |
| <1 day | 103,310 (82%) | 56,662 (83%) | 31,613 (85%) | 12,948 (74%) | 2,087 (78%) |
| ≥1 to <4 days | 14,344 (11%) | 6,619 (9.7%) | 4,391 (12%) | 2,904 (16%) | 430 (16%) |
| ≥4 to <7 days | 2,021 (1.6%) | 1,002 (1.5%) | 474 (1.3%) | 484 (2.7%) | 61 (2.3%) |
| ≥7 to <14 days | 2,341 (1.9%) | 1,419 (2.1%) | 309 (0.8%) | 562 (3.2%) | 51 (1.9%) |
| ≥14 days | 3,813 (3.0%) | 2,557 (3.7%) | 495 (1.3%) | 715 (4.1%) | 46 (1.7%) |
| Unknown | 15 | 15 | 0 | 0 | 0 |
| **Age at admission, median (IQR)** | **0 (0–0)** | **0 (0–0)** | **0 (0–0)** | **0 (0–1)** | **0 (0–0)** |
| Unknown | 15 | 15 | 0 | 0 | 0 |
| **Length of stay** |  |  |  |  |  |
| <1 day | 10,239 (8.1%) | 7,288 (11%) | 2,220 (6.0%) | 673 (3.8%) | 58 (2.2%) |
| ≥1 to <3 days | 35,318 (28%) | 26,064 (38%) | 5,892 (16%) | 3,216 (18%) | 146 (5.5%) |
| ≥3 to <7 days | 32,980 (26%) | 19,125 (28%) | 8,116 (22%) | 5,343 (30%) | 396 (15%) |
| ≥7 to <14 days | 21,089 (17%) | 9,074 (13%) | 7,242 (19%) | 4,012 (23%) | 761 (28%) |
| ≥14 days | 26,100 (21%) | 6,625 (9.7%) | 13,812 (37%) | 4,349 (25%) | 1,314 (49%) |
| Unknown | 118 | 98 | 0 | 20 | 0 |
| **Length of stay, median (IQR)** | **4 (2–11)** | **3 (1–6)** | **8 (3–21)** | **6 (3–13)** | **13 (7–24)** |
| Unknown | 118 | 98 | 0 | 20 | 0 |
| **Place of birth** |  |  |  |  |  |
| Inborn (born at this facility) | 93,865 (77%) | 52,352 (78%) | 27,858 (80%) | 11,671 (66%) | 1,984 (74%) |
| Outborn (born outside facility) | 28,089 (23%) | 14,567 (22%) | 6,928 (20%) | 5,903 (34%) | 691 (26%) |
| Unknown | 3,890 | 1,355 | 2,496 | 39 | 0 |
| **Condition at discharge** |  |  |  |  |  |
| Alive | 101,158 (81%) | 55,544 (82%) | 28,930 (78%) | 14,367 (82%) | 2,317 (87%) |
| Dead | 24,001 (19%) | 12,084 (18%) | 8,336 (22%) | 3,223 (18%) | 358 (13%) |
| Unknown | 685 | 646 | 16 | 23 | 0 |

<sup>1</sup> *n (%); Mean (SD); Median (Q1–Q3)*

Appendix 3: Primary Model Area Under Curve

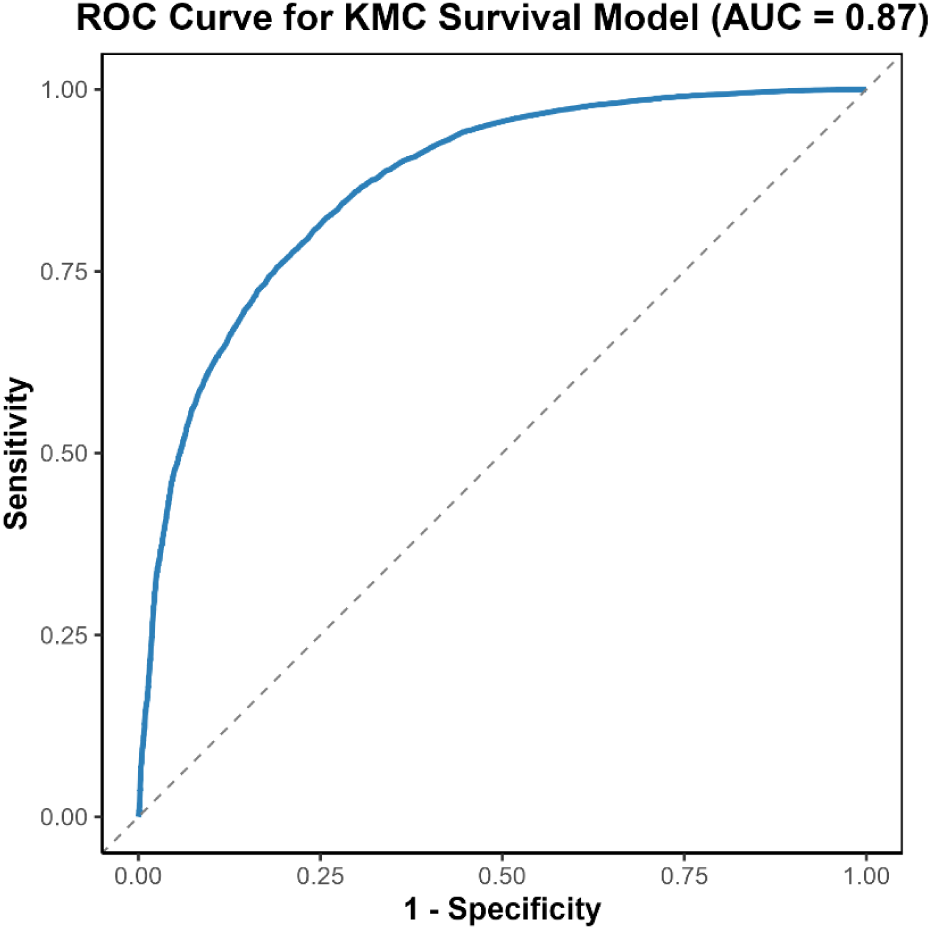

Appendix 4: Detailed Secondary Analysis of Timing of KMC Initiation

Detailed adjusted associations and birthweight-stratified predicted probabilities from the secondary analysis among KMC recipients are presented below. These estimates should be interpreted in the context of potential survivor selection bias, because neonates classified as having delayed or late KMC initiation necessarily survived to those later time points.

*Appendix Table 4.1: Adjusted associations for covariates (Mixed-effects Logistic Regression)*

| Covariate | Adjusted Odds Ratio (aOR) | 95% CI | p-value |
|---|---|---|---|
| Early KMC (1–3 days) vs Same-day KMC (≤24h) | 2.60 | (2.17, 3.11) | <0.001 |
| Delayed KMC (3–7 days) vs Same-day KMC | 3.69 | (2.97, 4.60) | <0.001 |
| Late KMC (>7 days) vs Same-day KMC | 7.41 | (5.80, 9.47) | <0.001 |
| Moderate severity (5–6) vs Mild (3–4) | 0.50 | (0.43, 0.58) | <0.001 |
| Severe severity (7–9) vs Mild (3–4) | 0.34 | (0.25, 0.48) | <0.001 |
| 1000–1499 g vs <1000 g | 4.23 | (3.40, 5.28) | <0.001 |
| 1500–1999 g vs <1000 g | 19.20 | (14.89, 24.77) | <0.001 |
| 2000–2499 g vs <1000 g | 53.91 | (38.69, 75.12) | <0.001 |
| Male vs Female | 0.70 | (0.61, 0.80) | <0.001 |
| Outborn vs Inborn | 0.83 | (0.71, 0.98) | 0.026 |

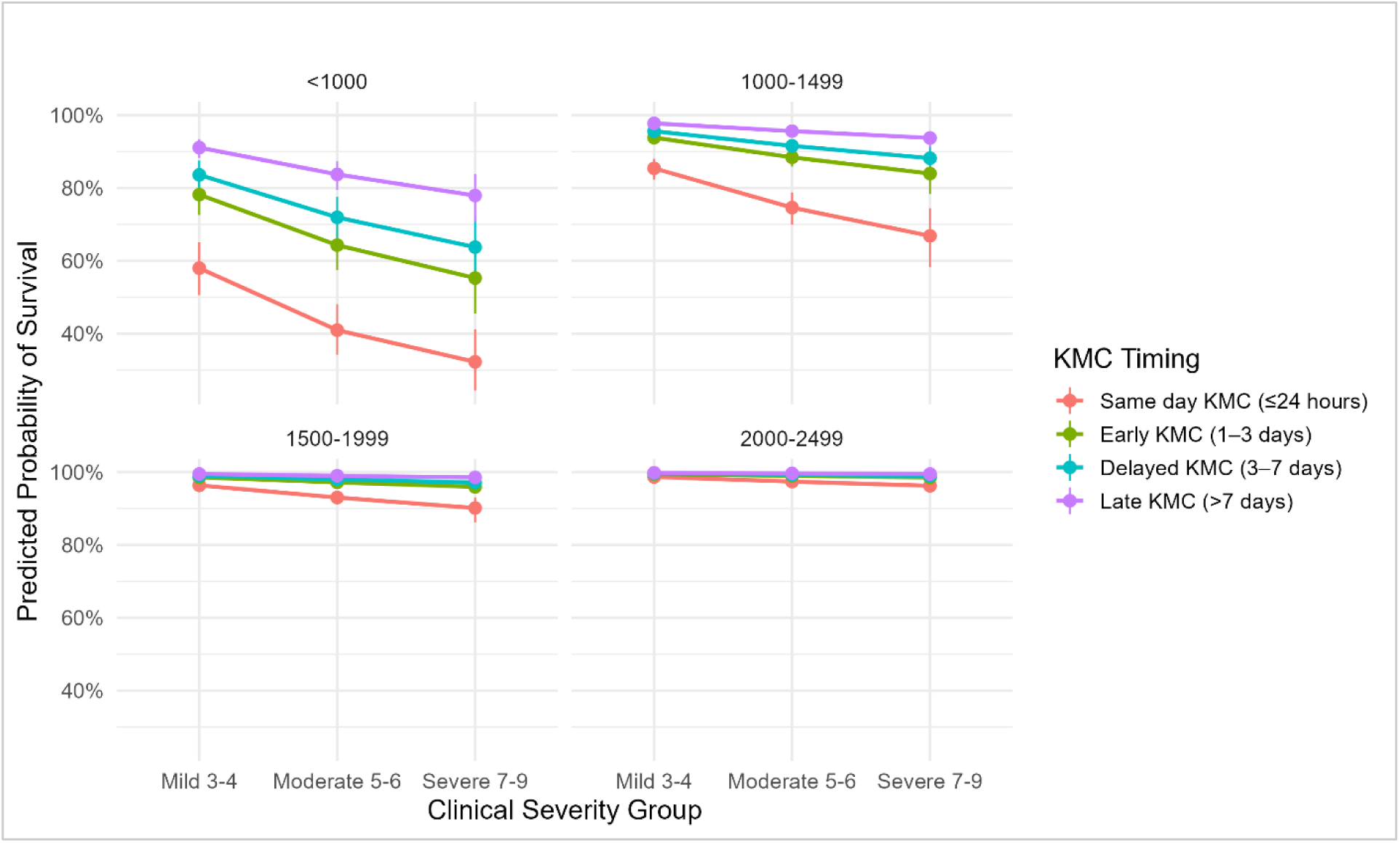

*Appendix Figure 4.1: Adjusted Predicted Survival by KMC Timing, Severity and Birthweight*

Appendix 5: Lay summary of associations between clinical factors and survival

| **What we looked at** | **What we found** |
|---|---|
| Birthweight under 1000 g | These babies are at by far the highest risk of dying and need the most intensive support |
| Birthweight 1000–1499 g | About 4 times more likely to survive than babies under 1000 g |
| Birthweight 1500–1999 g | About 19 times more likely to survive than babies under 1000 g |
| Birthweight 2000–2499 g | About 54 times more likely to survive than babies under 1000 g |
| Moderate illness at admission | Around half as likely to survive as babies who are only mildly ill |
| Severe illness at admission | Around a third as likely to survive as babies who are only mildly ill |
| Baby's sex | Boys were less likely to survive than girls |
| Born elsewhere and transferred in | Slightly less likely to survive than babies born in the same hospital |

Appendix 6: Ethical approvals for NEST360 implementation in Kenya, Malawi, Nigeria, and Tanzania

| **Country** | **Protocol Title** | **Protocol ID** |
|---|---|---|
| **Multi country (LSHTM)** | NEST360°: Complex evaluation of implementation for a health system package of hospital newborn care in Malawi, Kenya, Tanzania, and Nigeria | LSHTM: 21892 |
| **Kenya** | Using a Health Facility Assessment to Assess Quality of Newborn Care in Kenya | MSU/DRPI/MUERC/00810/19 |
|  | Evaluating the effects of technology and workforce enhancement to support neonatal hospital care in Kenya. | KEMRI/RES/7/3/1 |
|  | The Impact of NEST360 Interventions on Quality of Neonatal Care and Changes in Health Systems Resources on Neonatal Outcomes in Kenyan Health Facilities | MSU/DRPI/MUSERC/01200/23 |
|  | Applied Research to improve the quality of comprehensive newborn care through the introduction of the packages of the Newborn Essential Solutions and Technologies (NEST360) in Kenya | AKU/2025/ISERC-138 |
| **Malawi** | Using a Health Facility Assessment to Assess Quality of Newborn Care in Malawi | NHSRC 2463 |
| **Nigeria** | Quality Improvement Study of the Implementation of a Package of Trainings and Technologies for the Delivery of Comprehensive Newborn Care in Nigeria: A Multi-Country Study | LUTH: ADM/DCST/HREC/APP/3487 |
|  |  | UCH: UI/EC/20/0713 |
|  |  | NHREC: NHREC/01/01/2007 |
| **Tanzania** | Implementation study to improve the quality of comprehensive newborn care through introduction of the package of Newborn Essential Solutions and Technologies (NEST) in Tanzania | IHI: IHI/IRB/01-2021 |
|  |  | MUHAS: MUHAS-REC-12-2019-072 |
|  |  | NIMR: 3405 |
| **USA (Rice University)** | Using a Health Facility Assessment to Assess Quality of Newborn Care in Kenya | IRB-FY2020-266 |
|  | Quality Improvement Study of the Implementation of a Package of Trainings and Technologies for the Delivery of Comprehensive Newborn Care in Nigeria: A Multi-Country Study | IRB-FY2020-271 |
|  | Implementation study to improve the quality of comprehensive newborn care through introduction of the package of Newborn Essential Solutions and Technologies (NEST) in Tanzania | IRB-FY2020-237 |

Abbreviations: Maseno University Ethics Review Committee, NHSRC; Nigeria National Health Science Research Committee, LUTH; Lagos University Teaching Hospital, UCH; University College Hospital, NHREC; National Health Research Ethics Committee, IHI; Ifakara Health Institute, MUHAS; Muhimbili University of Health and Allied Science, NIMR; National Institute for Medical Research, IRB; Institutional Review Board
This table summarises ethical approvals obtained for the multi-country evaluation of the NEST360 small and sick newborn care package. Approvals were secured from institutional review boards and national ethics committees across Kenya, Malawi, Nigeria and Tanzania to cover data collection at the hospital and patient level.

Appendix 7: Neonatal clinical severity score for predicting survival

*Table 7.1: Severity Score for predicting survival*

| Variable | **Low Severity<br>(1 point per each category)** | **Moderate Severity<br>(2 points per each category)** | **High Severity<br>(3 points per each category)** |
|---|---|---|---|
| Gestational age | ≥32 weeks | ≥28 to <32 weeks | <28 weeks |
| SpO<sub>2</sub> on admission | 90–100% | 80–89% | <80% |
| Hypothermia on admission | ≥36.0–37.5°C (mild hypothermia or normal temperature) | 32.0–35.9°C | <32.0°C |
| Total score | Severity category |  |  |
| 3–4 | Mild (low) |  |  |
| 5–6 | Moderate |  |  |
| 7–9 | Severe (high) |  |  |

Note: SpO<sub>2</sub> = peripheral oxygen saturation; °C = degrees Celsius. Component thresholds
Composite score: Sum of the three component scores (Lowest possible score = 3; highest possible score = 9)

